# Deep Learning Based Cardiac Cine Segmentation – Transfer Learning Application to 7T Ultrahigh-Field MRI

**DOI:** 10.1101/2020.06.15.20131656

**Authors:** Markus Johannes Ankenbrand, David Lohr, Wiebke Schlötelburg, Theresa Reiter, Tobias Wech, Laura Maria Schreiber

## Abstract

**Background:** Artificial neural networks show promising performance in automatic segmentation of cardiac MRI. However, training requires large amounts of annotated data and generalization to different vendors, field strengths, sequence parameters, and pathologies is limited. Transfer learning addresses this challenge, but specific recommendations regarding type and amount of data required is lacking. In this study we assess data requirements for transfer learning to experimental cardiac MRI at 7T where the segmentation task can be challenging. In addition, we provide guidelines, tools, and annotated data to enable transfer learning approaches by other researchers and clinicians.

**Methods:** A publicly available segmentation model was used to annotate a publicly available data set. This labelled data set was subsequently used to train a neural network for segmentation of left ventricle and myocardium in cardiac cine MRI. The network is used as starting point for transfer learning to 7T cine data of healthy volunteers (n=22; 7873 images). Structured and random data subsets of different sizes were used to systematically assess data requirements for successful transfer learning.

**Results:** Inconsistencies in the publically available data set were corrected, labels created, and a neural network trained. On 7T cardiac cine images the initial model achieved DICE_LV_=0.835 and DICE_MY_=0.670. Transfer learning using 7T cine data and ImageNet weight initialization improved model performance to DICE_LV_=0.900 and DICE_MY_=0.791. Using only end-systolic and end-diastolic images reduced training data by 90%, with no negative impact on segmentation performance (DICE_LV_=0.908, DICE_MY_=0.805).

**Conclusions:** This work demonstrates and quantifies the benefits of transfer learning for cardiac cine image segmentation. We provide practical guidelines for researchers planning transfer learning projects in cardiac MRI and make data, models and code publicly available.

## Background

Image segmentation, which is of great interest in cardiac magnetic resonance imaging is applied to partition acquired images into functionally meaningful regions, allowing the extraction of quantitative static measures such as myocardial mass, left ventricle (LV) volume, right ventricle (RV) volume, and wall thickness, as well as dynamic measures such as wall motion and the ejection fraction (EF). Cardiac cine MRI is the accepted gold standard for this assessment of cardiac function^1^ and anatomy and is therefore of paramount clinical importance^2,3^. Proper segmentation of such data sets is a tedious and time-consuming process that has increasingly been tackled using various deep learning approaches^4-7^.

Artificial neural networks have been shown to outperform other methods on several high profile image analysis benchmarks and, thus, so-called deep learning models have become state-of-the-art for a wide variety of computer vision tasks. Multiple factors like the wide application area of deep learning, available compute power, and increasing investments as well as user-friendly open source software have enabled a rapid development of the field of artificial intelligence. This led to ever increasing applications in medical imaging such as MRI^8^ where tasks nowadays range from data acquisition and image reconstruction^9-11^, image restoration^12,13^, to image registration^14,15^, segmentation^16-19^ as well as classification^20,21^ and outcome prediction^22,23^.

There is consensus in the field that the limited availability of labelled or annotated data due to data access, privacy issues, missing data harmonization, and data protection is one of the main obstacles for future clinical applications of deep neural networks^17,19,24^. While some resources like the UK Biobank^25^ already exist to address this issue, the high quality standards and the amount of work required to organize and maintain such a resource makes data access expensive. In addition, such data may already exceed the quality that is available in clinical routine cardiac MRI. This leads to neural networks, which perform very well for a very specific task within a confined data space, where training and testing data share the same distribution. However, these networks usually lack generalization capabilities. While methods such as data augmentation, transfer learning, weakly-, self-supervised, and unsupervised learning have been applied to overcome the issue of small datasets in research, it is unclear how much data is really required in order to create a well-generalizing network or to apply transfer learning.

In this work, we aim to enable researchers and clinicians in cardiology to apply deep learning-based segmentation models in their respective research by providing guidelines and easily accessible tools as well as annotated data for transfer learning. We create labels for a public data set, the Data Science Bowl Cardiac Challenge Data^26^ (further referred to as Kaggle data set) which, at this point, does not have segmentation labels. We further create a base network for LV segmentation using these labels and evaluate its performance on 7T human cine data. In addition, we assess if transfer learning improves model performance for the 7T segmentation task and analyze how much and which data is required. The framework provided in this study in combination with access to scripts and the data used, will enable researchers to reproduce our results and apply deep learning based segmentation in their respective field.

## Methods

### The Kaggle Data Set

As mentioned above, cardiac MRI is the gold standard for the assessment of cardiac function, a key indicator of cardiac disease. The 2015 Data Science Bowl challenged participants to create an algorithm for automatic assessment of end-systolic and end-diastolic volumes (ESV and EDV) and thus, ejection fraction, based on cardiac cine MRI. The data set consists of a training, a validation, and a test set and once the challenge has ended, all sets and their corresponding volume information (end-systolic and end-diastolic) was made available for research and academic pursuits, leading to a total of 1140 “annotated” cardiac MRI examinations of normal and abnormal cardiac function. Images are in DICOM format resolving up to 30 phases of the cardiac cycle. While we will focus on short axis images in this study, the Kaggle data set also contains alternative views. Examinations were done on 1.5 T and 3.0 T systems (Siemens Magnetom Aera and Skyra, Siemens Healthineers, Erlangen, Germany) with applications of both FLASH and TrueFISP sequences. An overview of the complete data set and its variation in patient data and sequence parameters is given in Table 1.

**Table 1:**
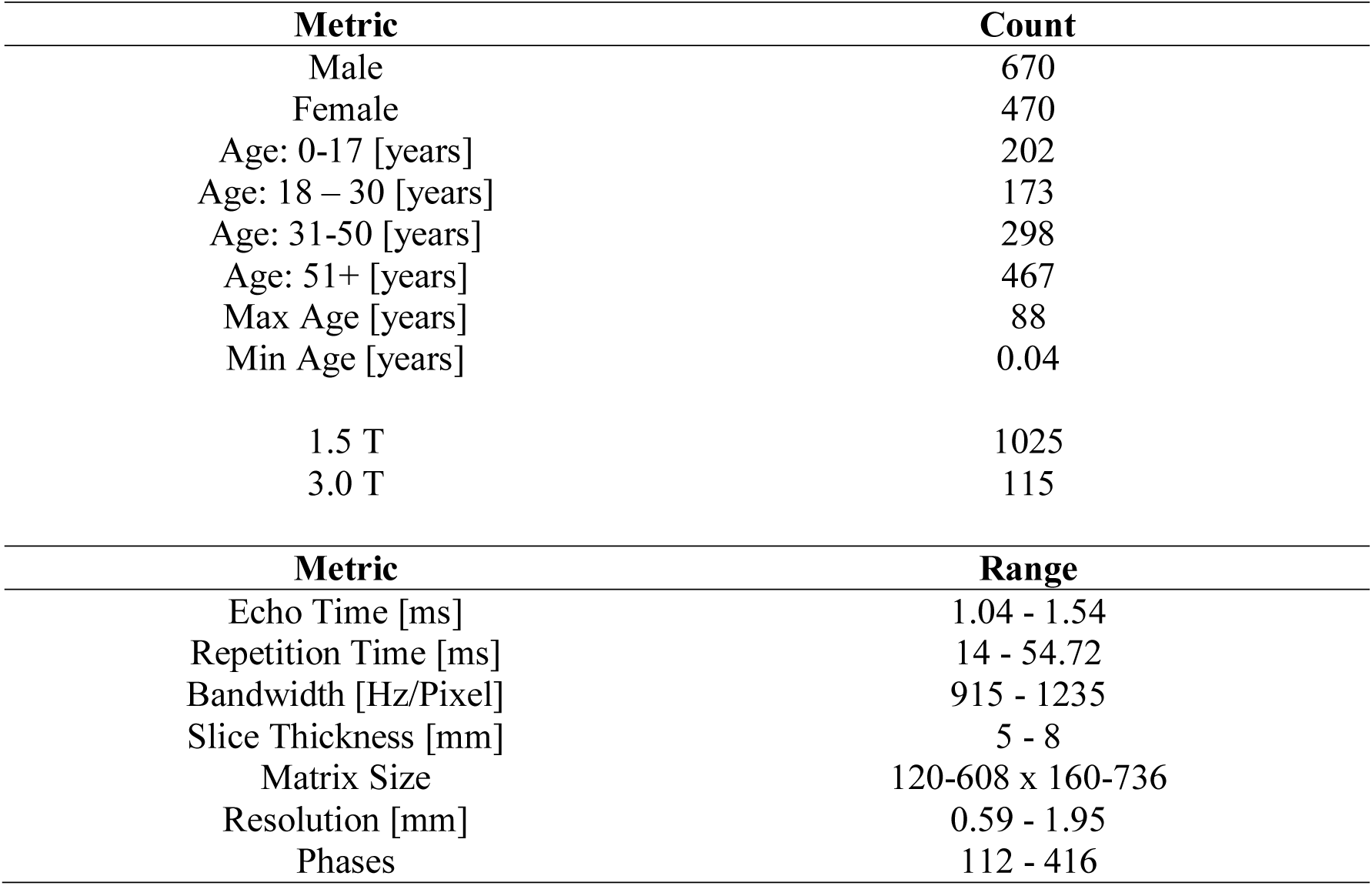
Data Composition and measurement parameters of the Kaggle data

| **Metric** | **Count** |
| --- | --- |
| Male | 670 |
| Female | 470 |
| Age: 0-17 [years] | 202 |
| Age: 18 – 30 [years] | 173 |
| Age: 31-50 [years] | 298 |
| Age: 51+ [years] | 467 |
| Max Age [years] | 88 |
| Min Age [years] | 0.04 |
| 1.5 T | 1025 |
| 3.0 T | 115 |
| **Metric** | **Range** |
| Echo Time [ms] | 1.04 - 1.54 |
| Repetition Time [ms] | 14 - 54.72 |
| Bandwidth [Hz/Pixel] | 915 - 1235 |
| Slice Thickness [mm] | 5 - 8 |
| Matrix Size | 120-608 x 160-736 |
| Resolution [mm] | 0.59 - 1.95 |
| Phases | 112 - 416 |

### Data Curation

The complete data set is a compilation of real, clinical data from several sites and as such, subject to inconsistencies within individual examinations. Those can be a combination of:

▪ missing time points
▪ inconsistent slice spacing
▪ inconsistent image dimension
▪ repeat measurements (identical slice location)
▪ scaled images
▪ image rotations

Prior to the application of the published segmentation network of Bai et al.^4^ we performed data curation, correcting inconsistencies in all but 8 examinations. More detailed information and curated data can be found in the online repository (https://github.com/chfc-cmi/cmr-seg-tl, https://doi.org/10.5281/zenodo.3876351).

### Creating Labels

Once the data was corrected for inconsistencies we ran the Python based segmentation model of Bai et al. for the complete data set, generating RV, LV, and blood pool labels as well as LV ESV and EDV volumes. ESV and EDV values were then compared to the ground truth values provided by Kaggle in order to determine the accuracy of the network prediction. Based on this comparison we created confidence sets where the predicted values were in the range of ±5% (p5), ±10% (p10), and ±15% (p15) of the true value. Respectively, these sets contained 175, 520, and 763 examinations and 54540, 162480, and 238350 images. All scores (label versus ground truth) for ESV and EDV values are listed in the online repository.

### Hardware

In order to deal with the extensive computation demands we used a custom workstation and a high performance cluster, both with graphical processing units. Details are given in the online repository.

### Framework - Deep neural network

All implementations were realized using Pytorch^27^ and fastai^28^ V1. Training of neural networks (U-Net^29^ architectures with varying backbones: Restnet34^30^, ResNet50^31^, and VGG16^32^) was performed using fastai’s implementation of the *one cycle* policy^33^ with adjusted learning rates (lr) and the confidence sets p5, p10, and p15.

### Parameter Search

During the parameter search, we evaluated the influence of different training parameters on the efficacy of the trained model. Training speeds of the varying models and their architectures is given in the online repository. Training with a weight-decay of 0.02 and a batch-size of 32 was done for 30 epochs with frozen weights (lr = 1e^-4^) and another 30 epochs with unfrozen weights (lr = 1e^-5^). Details regarding frozen and unfrozen weights are provided in the online repository. The smallest training set (p5) was used initially, image size was 256×256, and moderate data augmentation transforms (s1: flip [none], rotation [20°], lighting [0.4], zoom [1.2], padding [zeros]) were applied.

In order to avoid an extensive parameter grid search, we assessed parameter dependent performance changes in incremental steps. After each step, we determined the best-performing model using EF predictions and introduced subsequent parameter variations on this respective model.

In the first step we evaluated the influence of the architecture (VGG16, ResNet34, ResNet50) compared to the fully convolutional Network by Bai et al^4^. trained on UKBB data (further referred to as UKBB model). Due to memory limitations, we had to reduce the batch size for training of the VGG16 and the ResNet50 models.

In the second step, we assessed variations in the loss function such as cross-entropy (default), generalized DICE^34^, and focal loss. In the third and last step we evaluated the influence of the number of training images using the confidence sets p5, p10, and p15.

We assessed the influence of training data resolution, training a model with lower input resolution (128×128, r34_CE_p5_128). Details are provided in the online repository.

### Data Augmentation

Since transfer learning applications assessed in this study are based on 7T data we expect somewhat different image contrast and artefacts compared to conventional, clinical datasets. In addition, we intended to account for the heterogeneous training data, which led to the following set of augmentations for the initial networks (s1: flip [none], rotation [20°], lighting [0.4], zoom [1.2], padding [zeros]). Further, we aimed to introduce some robustness to forms of data variations, such as 90°-rotations and flips (left-right) using more extensive data augmentation (s2: flip [Left-Right], rotation [90°], lighting [0.4], zoom [1.2], padding [zeros]). In order to test the efficacy of these transforms we trained a new model (r34_CE_p5_s2) and compared EF predictions on a dataset including rotated and flipped images retained during the data curation process.

### Transfer Learning

All assessments regarding transfer learning to 7T data are done using model: r34_CE_p5_s2. As initial point of comparison we used the UKBB model to create labels for 7T data, in order to assess generalization capability of a model, which was trained on a very homogeneous data set (UKBB).

Following approval of the local ethics committee (7/17-SC), n=22 (14 female, 8 male) were examined using a 7T whole body MRI system (Siemens MAGNETOM Terra, Erlangen, Germany) and a 1TX/16RX thorax coil (MRI Tools, Berlin, Germany)^35^. Written informed consent was obtained prior to all measurements. Patient age was 22-53 years, body weight 52-95 kg, and height: 151-185cm. For triggering, both the integrated ECG and an external acoustic triggering system (MRI Tools, Berlin, Germany) were used in order to synchronize measurements with the heartbeat, choosing whichever method provided a more stable trigger signal during the examination. Images were obtained during initial sequence implementation and optimization for 7T cardiac MRI using a cardiovascular (CV) GRE cine-sequence and protocol parameters therefore vary to some degree. The parameters were: TE = 3.57ms, FOV = 340 mm x 320 mm, interpolated voxel size = 0.66 x 0.66 x 6 mm, GRAPPA acceleration factors: R = 2 and R = 3. Depending on the heart rate 6-11 segments and 20-35 cardiac phases were measured using retrospective gating. Short axis CINE stacks for volumetric evaluation varied in the number of slices (14-17) and multiple breath-holds (∼13s) were necessary to acquire the whole stack. Images were assigned into training, validation and test sets (14, 5, 3 subjects and 5076, 1842, 955 images, respectively). All images were manually segmented by an expert radiologist (TR). Three data sets of the test set were additionally segmented by an expert cardiologist (WS), in order to obtain an estimate of interobserver-variability.

### Starting Point for Model Training - 7T Human

To assess the efficacy of transfer learning for LV segmentation based on clinical 1.5T and 3T data and experimental (human) 7T data, we compare models with varying degrees of training and transfer learning. Using a U-Net architecture with a ResNet34 backbone (r34_CE_p5_s2), we generated the following three models:

1. initialization with random weights (R)
2. initialization with ImageNet-weights – transfer learning 1 (TL)
3. Model 2, pre-trained on Kaggle data - transfer learning 2 (TL^2^)

All models were used to generate predictions for the 7T test set. Model performance was always evaluated using the Soerensen-DICE^36^ coefficient between predictions and respective ground truth labels.

### Data Requirements for Model Training - 7T Human

To assess how much and what data is required for convergence of a model we trained all models (R, TL, TL^2^) with subsets of the training data. These subsets were created in two ways:

1. Complete subject data (all slices and all phases) from either 14, 7, 3, 1 subjects (5076, 2626, 1001, 306 images, respectively); Partial subject data (only end-systolic and end-diastolic images) from all subjects (448 images)
2. Shuffle all images once, create a list of images (1-5076), and generate subsets corresponding to the respective image numbers from subset 1, always starting the count with image #1

When training with subsets, the model is exposed to a smaller number of images in every epoch. We therefore increased the number of epochs for the subsets to correct for this effect.

## Results

### Framework - Deep neural network

#### Parameter Search

Results of the parameter search are illustrated in Figure 1, showing the absolute distance between the EF predictions based on model segmentation and ground truth data provided by Kaggle. Overall, the impact of parameter variation on model performance was small (3.64-4.06% mean distance to ground truth EF).

**Figure 1:**
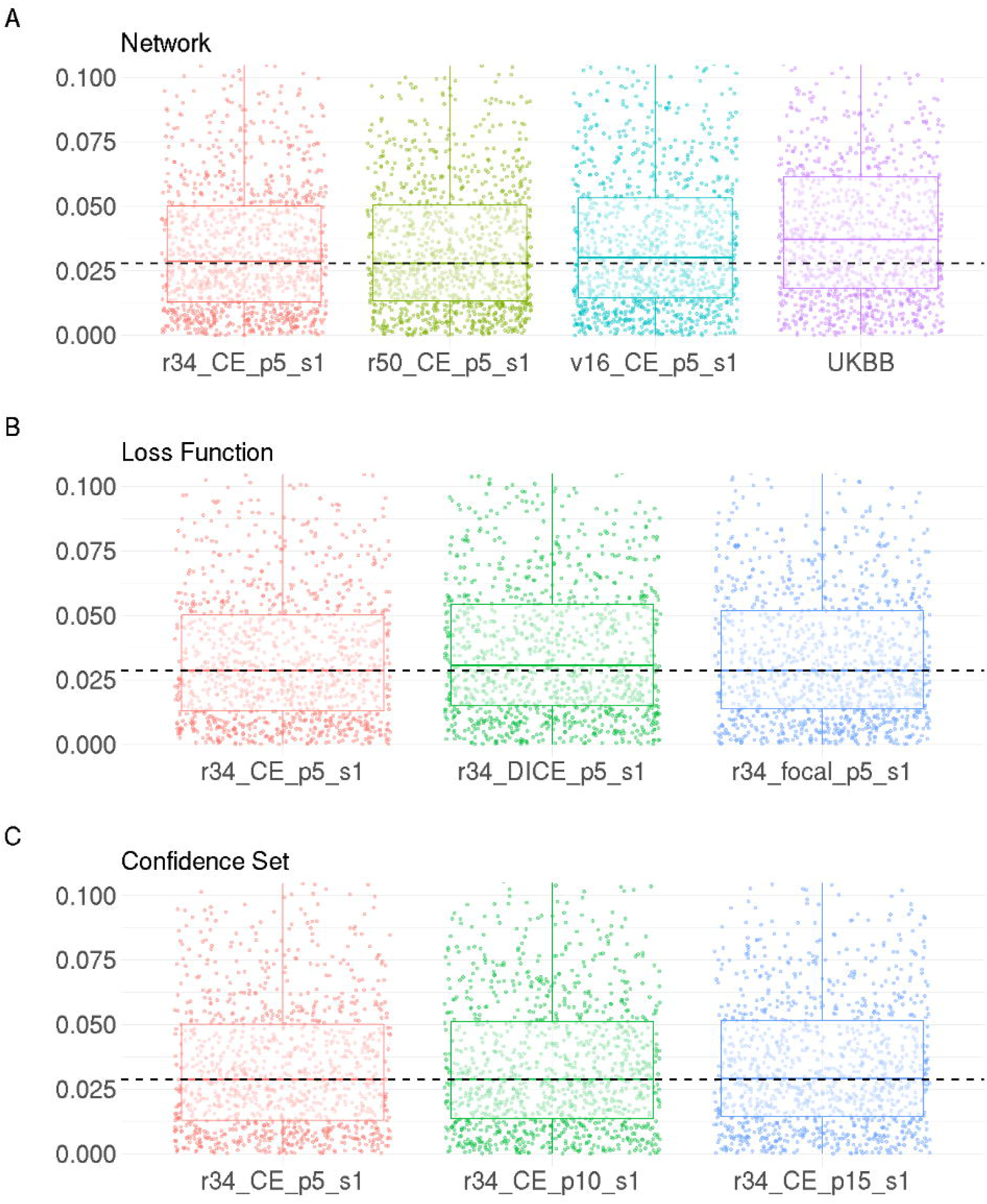
Model evaluation during incremental parameter search. Plots show the absolute distance between the EF prediction based on model segmentation and ground truth data provided by Kaggle. The range of the y-axis is restricted for better comparability, dashed lines indicates lowest median. Model performance with **A:** Architectures (r34: ResNet34, r50: ResNet50, VGG16: v16, UKBB). **B:** Loss functions (Cross-entropy: CE, DICE, focal loss). **C:** Confidence sets (p5: 5%, p10: 10%, p15: 15%).

In a first approach to interpret these results, we compared varying architectures, such as ResNet34, ResNet50, and VGG16 with the UKBB model (Figure 1A). All models led to lower mean and median distance values compared to the UKBB model (table 2). The lowest median distance values were found using a ResNet50 (2.79%), while the lowest mean distance values were found using a ResNet34 (3.64%). Differences in the absolute distance between the models (r34, r50) were rather small (Δ0.08%), however. Considering computational demand, we selected the ResNet34.

**Table 2:**
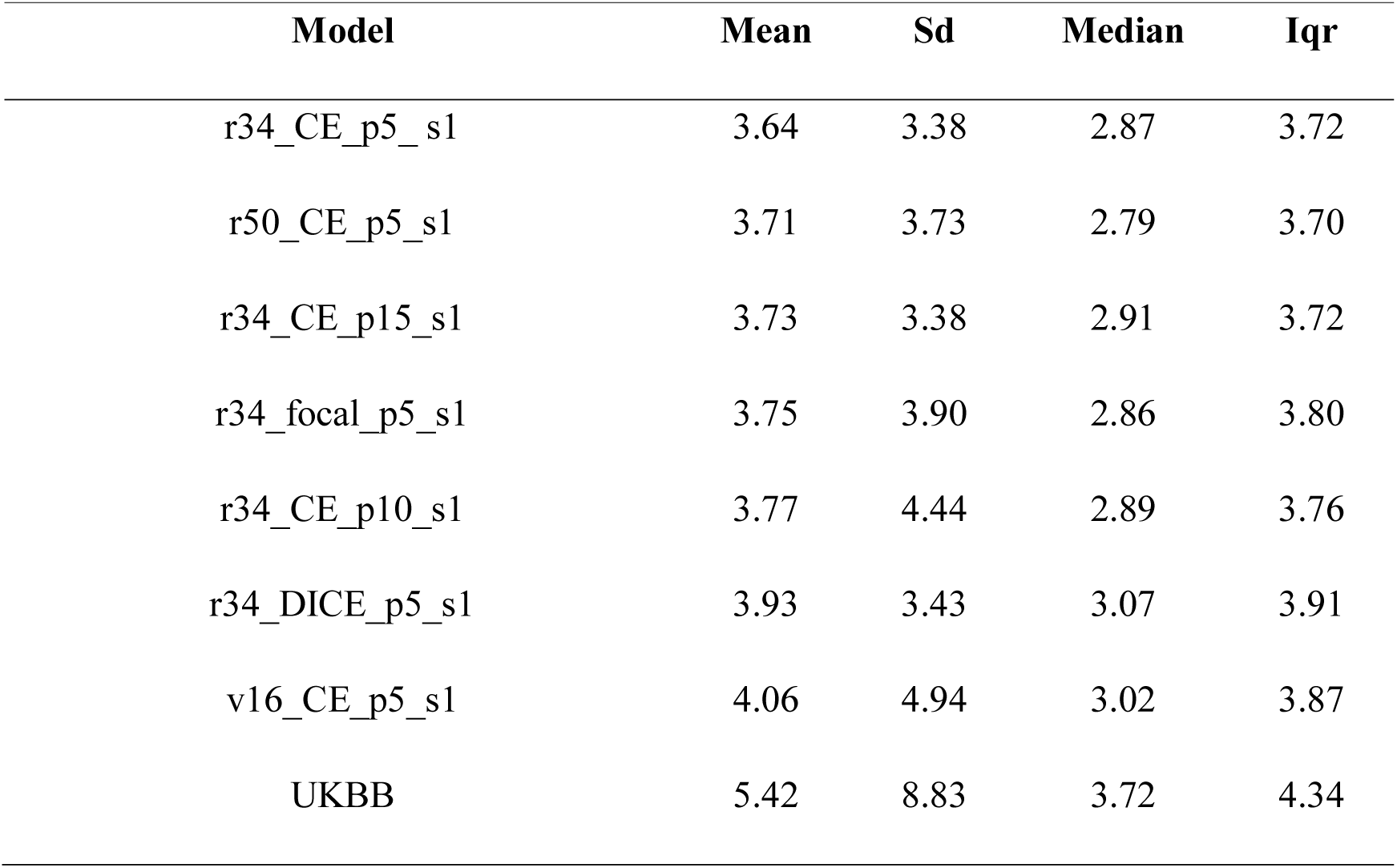
Summary statistics of absolute deviation of predicted and true EF in % for the parameter search. Sorted from lowest to highest mean value. Models are named by architecture (ResNet34: r34, ResNet50: r50, VGG16: v16), loss function (cross entropy: CE, focal, DICE), confidence set (p5, p10, p15), and data augmentation (s1, s2).

| Model | Mean | Sd | Median | Iqr |
| --- | --- | --- | --- | --- |
| r34_CE_p5_s1 | 3.64 | 3.38 | 2.87 | 3.72 |
| r50_CE_p5_s1 | 3.71 | 3.73 | 2.79 | 3.70 |
| r34_CE_p15_s1 | 3.73 | 3.38 | 2.91 | 3.72 |
| r34_focal_p5_s1 | 3.75 | 3.90 | 2.86 | 3.80 |
| r34_CE_p10_s1 | 3.77 | 4.44 | 2.89 | 3.76 |
| r34_DICE_p5_s1 | 3.93 | 3.43 | 3.07 | 3.91 |
| v16_CE_p5_s1 | 4.06 | 4.94 | 3.02 | 3.87 |
| UKBB | 5.42 | 8.83 | 3.72 | 4.34 |

In the next step of the parameter search we evaluated model performance using varying loss functions, namely cross-entropy, generalized DICE, and focal loss (Figure 1B). Using the generalized DICE score led to the highest mean (3.93%) and median (3.07%) distance values. Median distance values were similar for cross-entropy and focal loss (2.87% vs 2.86%), while the mean distance value was lowest using cross-entropy (3.64%).

We thus selected cross-entropy for the next step of the parameter search, where we evaluated model performance using varying confidence sets: 5%, 10%, 15% (Figure 1C). Using the various confidence sets only slightly affected median distance values (2.87%, 2.89%, 2.91%). Based on EF predictions the model: r34_CE_p5_s1 performed best achieving a mean distance value of 3.64%.

### Data Augmentation

Figure 2 shows the performance of our models on the image set containing rotated images, plus the performance of an additional model where data augmentation allowed left-right flips, as well as rotations of up to 90°. Median and mean absolute distance values were lowest (3.06%, 4.08%) using the model with extended data augmentation (r34_CE_p5_s2).

**Figure 2:**
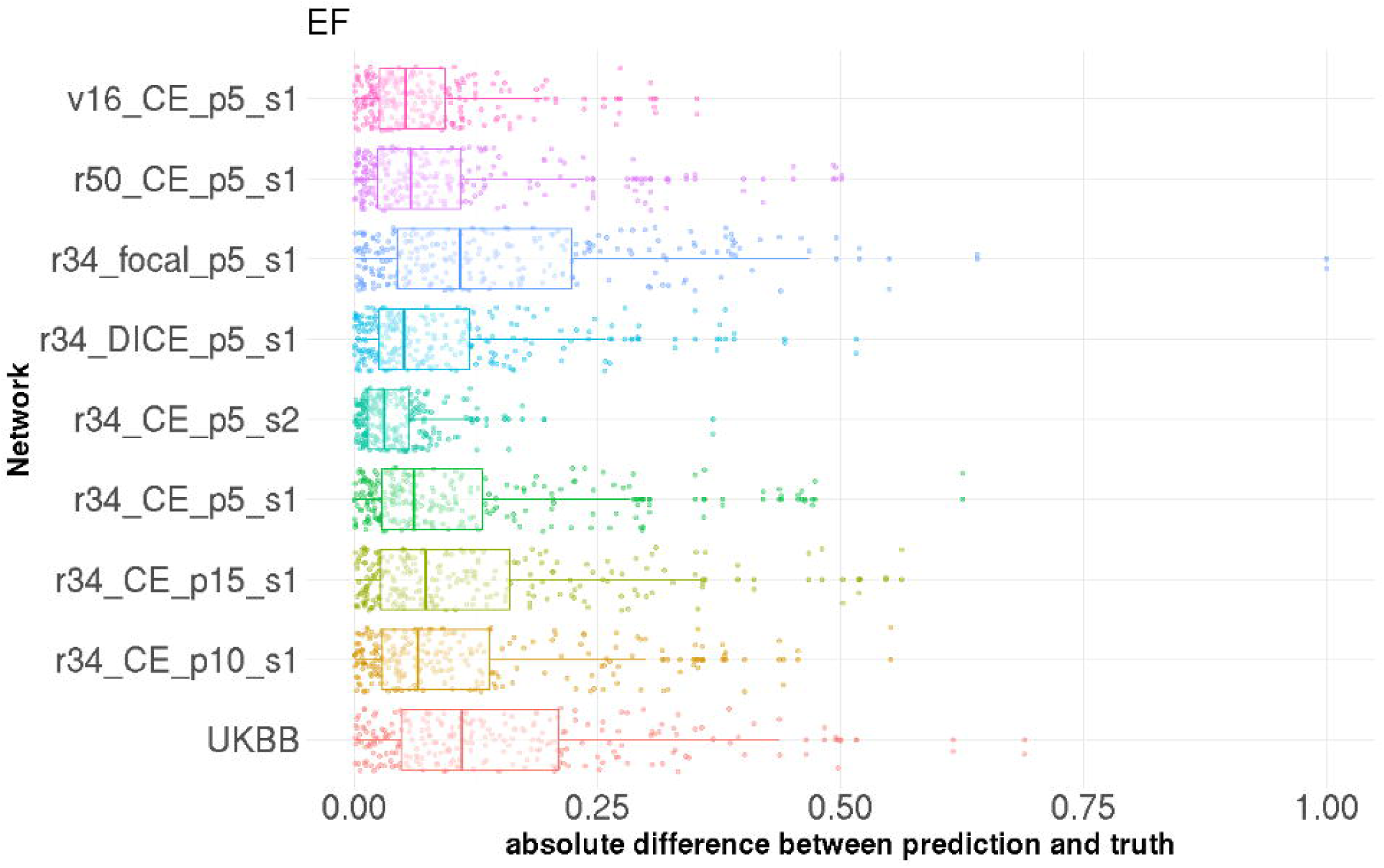
Model evaluation based on data including rotated images. Plots show the absolute distance between the EF predictions based on model segmentation and ground truth data provided by Kaggle for all models of the parameter search, plus one model trained with extended data augmentation (s2). Models are named by architecture (ResNet34: r34, ResNet50: r50, VGG16: v16), loss function (cross entropy: CE, focal, DICE), confidence set (p5, p10, p15), and data augmentation (s1: standard data augmentation, s2: extended data augmentation, enabling LR-flips and rotations up to 90°).

**Figure 3:**
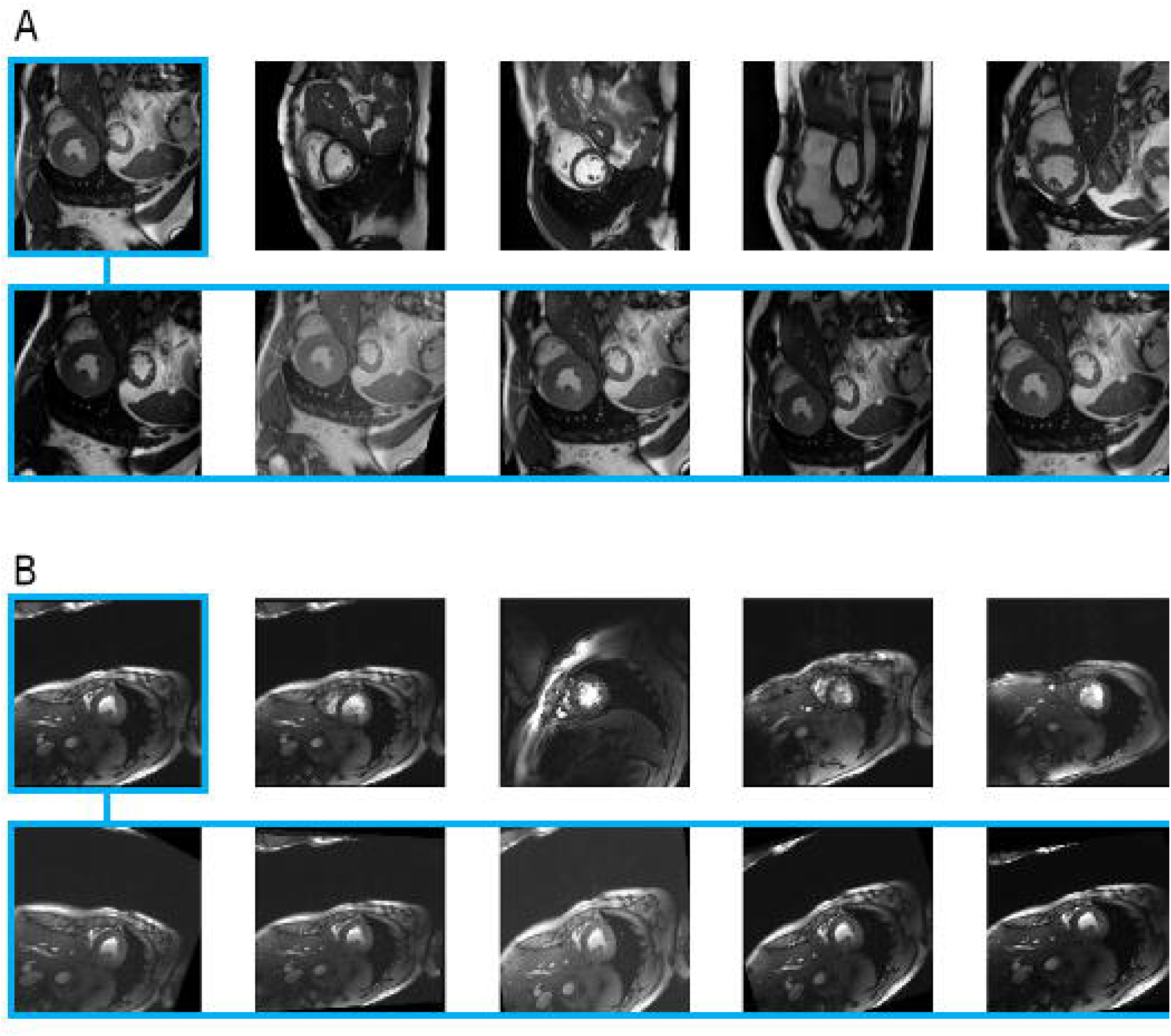
Exemplary cine images and respective data augmentation. Random selection of five images (top) with five data augmentation examples (bottom) for the first image of the random selection. A: Kaggle data. B: 7T human cine data.

### Transfer Learning

Exemplary cine images from the Kaggle and the 7T cine data set as well as respective data augmentation are shown in **Error! Reference source not found**.A and 3B. While the Kaggle data set includes images with varying field of views and resolution, the 7T data is consistent.

Figure 4 presents the inter-observer variability as difference of LV volume within each image in ml for all slices and phases of the 7T human cine test set (n=3). The slice count starts with 0 at the most apical slice and moves towards the most basal slice with increasing slice number. Overall expert 2 achieved DICE_LV_ = 0.94 and DICE_MY_ = 0.81 and deviations in LV volume of individual images were lower than ±5 ml in all but one image (set 3, slice 12, phase 4). Compared to expert 1, who labelled our training data, and expert 2 the AI model achieved DICE_LV_ = 0.90, DICE_MY_ = 0.79 as well as DICE_LV_ = 0.91, DICE_MY_ = 0.81. Deviations in LV volume of individual images were smaller than 5 ml in >95% of the cases. Exemplary predictions of the AI and deviations to expert 1 are shown in Figure 5. All apical slices labelled by the AI were in excellent agreement with that of our experts. The largest deviations between AI and both experts was found for the very basal slice where myocardial tissue moves in and out of plane throughout the cardiac cycle.

**Figure 4:**
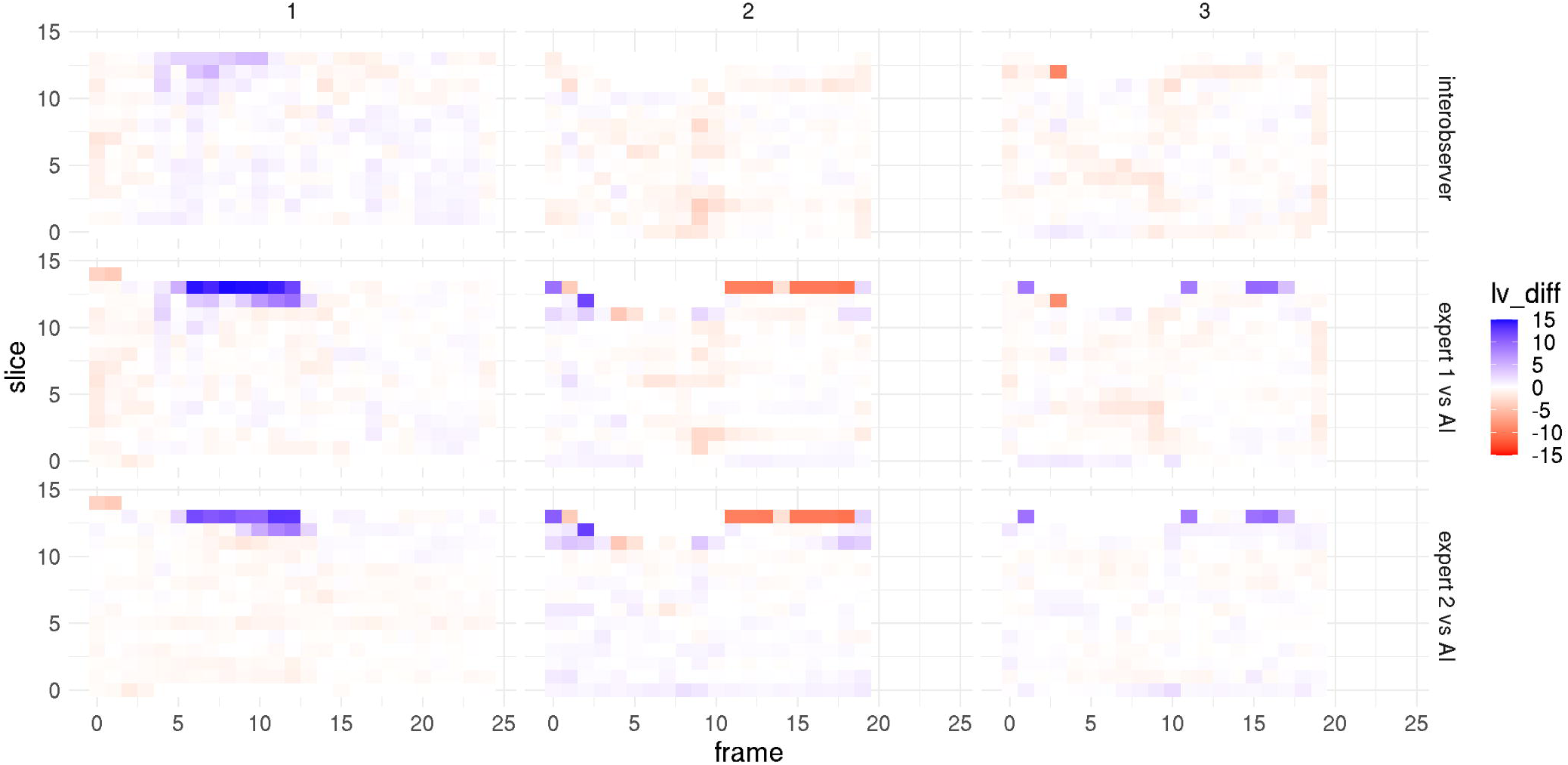
Inter-observer variability. Difference in LV volume in [ml] for all slices and phases of the 7T cine images of the test set. The slice count starts with 0 at the most apical slice and moves towards the most basal slice with increasing slice number. Top: Inter-observer variability of the two experts. Middle: Inter-observer variability expert 1 (labelled training data as well) versus AI. Bottom: Inter-observer variability expert 2 versus AI.

**Figure 5:**
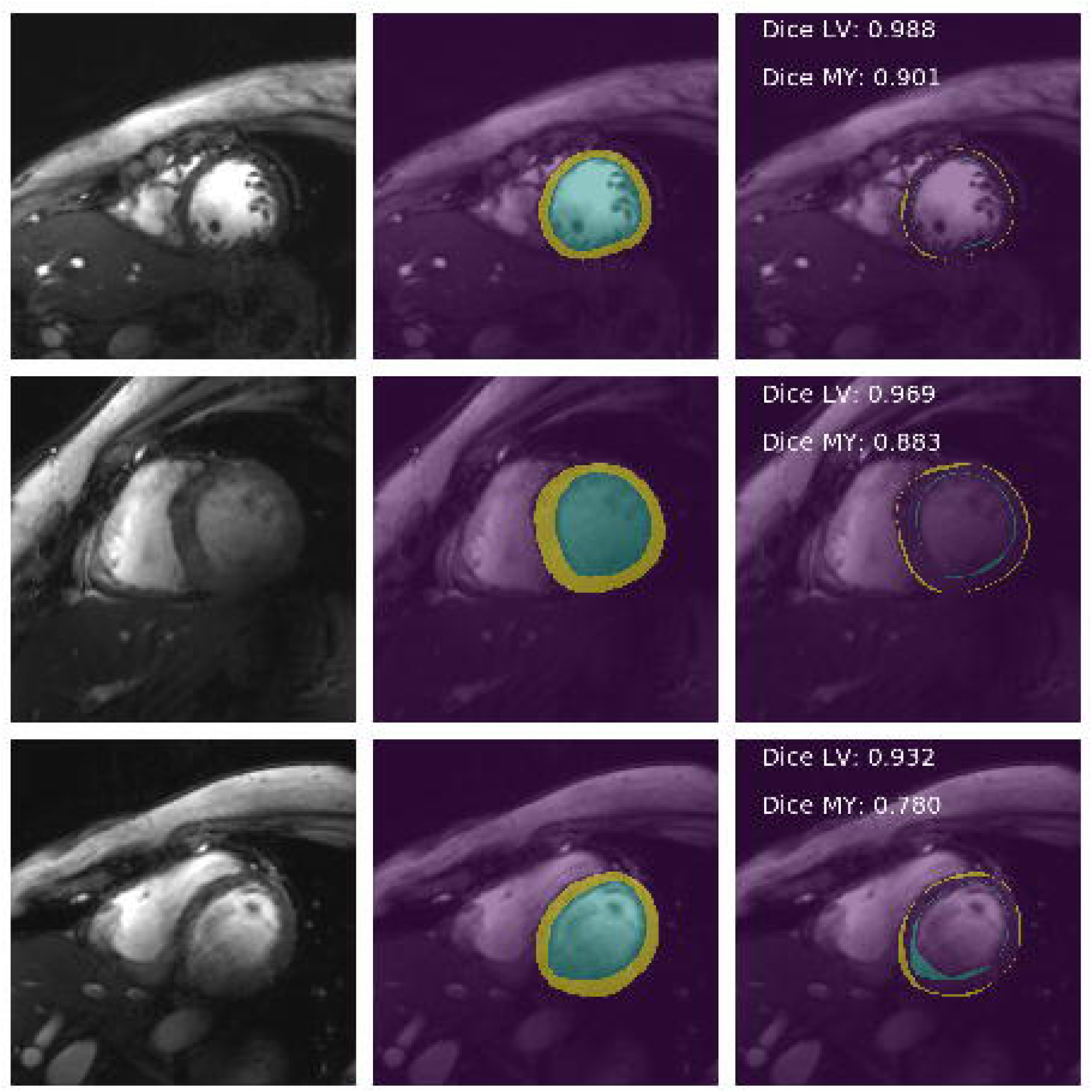
Predictions of TL^2^ on the 7T human test set. Examples of mid-cavity segmentation results with high (top), intermediate (middle) and low (bottom) DICE scores. Images (left), with predicted classes (middle, background: purple, LV: blue, MY: yellow) and differences to the ground truth (right, LV-error: blue, MY-error: yellow)

### Starting Point for Model Training

Results of model training using varying degrees of transfer learning are displayed in Figure 6. Plotted are the DICE scores for the left ventricle and the myocardium in dependence of the number of images seen during training, showing performance and overall convergence for the three models analyzed. All curves have been smoothed to increase interpretability. Respective plots of the raw data are shown in the online repository.

**Figure 6:**
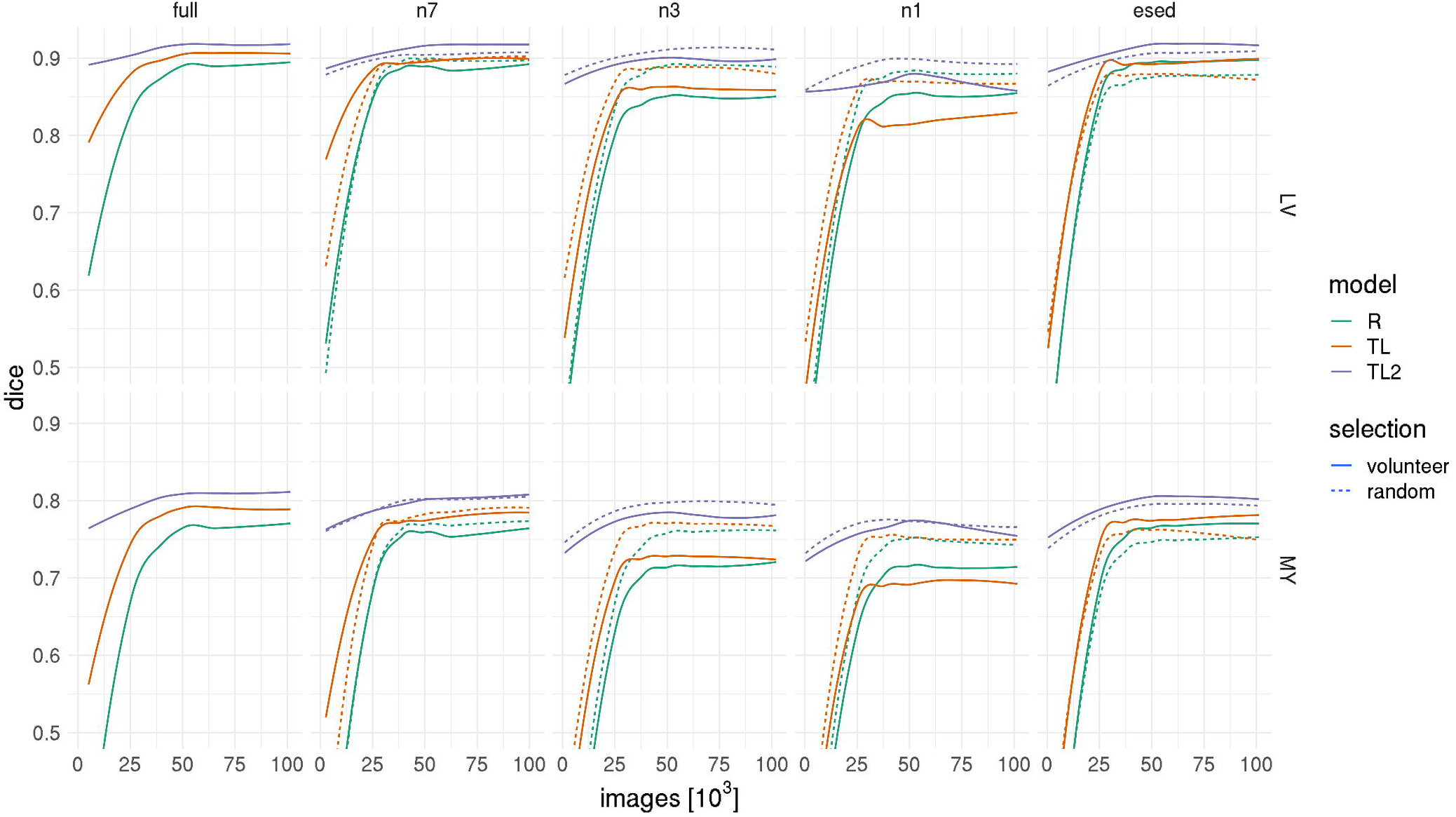
Training evaluation based on the validation set. DICE scores of the left ventricle and the myocardium in 7T human cardiac cine images normalized for the number of images seen. Varying degrees of transfer learning (R: ResNet34 initialized with random weights and trained using 7T cine images, TL: ResNet34 initialized with ImageNet weights and trained using 7T cine images, TL^2^: ResNet34 initialized with ImageNet weights, pre-trained on the 1.5T and 3T Kaggle cine images and re-trained on 7T cine images) are shown for the two subsets 1 (line): subset of whole volunteers (full=14, 7, 3, 1), 2 (dotted line): subset of random images with image numbers corresponding to first subset. In addition, there is one model (“esed”) trained using only end-systolic and end-diastolic images from all volunteers and a corresponding model trained with a number of random images equivalent to the “esed”-set.

Starting with the full data set, there are clear differences in starting points (DICE after first epoch), convergence speed, and peak performance (highest performance reached) for the three models.

R: Random weight initialization followed by training using 7T data led to the:

- lowest starting points with DICE_LV_ ∼ 0.57 and DICE_MY_ ∼ 0.25
- lowest convergence speed, reaching plateau values after 100.000 images
- lowest peak performance with DICE_LV_ ∼ 0.89 and DICE_MY_ ∼0.77

TL: ImageNet weight initialization followed by training using 7T data led to the:

- starting points of DICE_LV_ ∼ 0.77 and DICE_MY_ ∼ 0.51
- higher convergence speed, reaching plateau values after 50.000-60.000 images
- higher peak performance with DICE_LV:_ 0.91 and DICE_MY_: 0.79

TL^2^: ImageNet weight initialization, pre-trained (Kaggle data), re-trained 7T data led to the:

- highest starting points with DICE_LV_ ∼ 0.90 and DICE_MY_ ∼ 0.78
- higher convergence speed, reaching plateau values after 40.000-50.000 images
- higher peak performance with DICE_LV:_ 0.92 and DICE_MY_: 0.81

### Data Requirements for Model Training

Results of model training using varying degrees of transfer learning and a smaller amount of training data are displayed in Figure 6 as well. The full training data set consists of 14 volunteers, while the subsets consist of 7, 3, and 1 volunteer. For the most part, curves follow the trend described for the full data set, while each reduction in volunteers led to lower starting points. Peak performances remain similar with a reduction to 7 volunteers, but drop using subset n3, in particular for models R and TL. Only for a very small number of training images (n1) peak performances are higher for model R (DICE_LV_: 0.86; DICE_MY_: 0.72) compared to TL (DICE_LV_: 0.83; DICE_MY_: 0.70).

For small subsets, such as n3 and n1, starting points as well as peak performances of all models is higher using the random selection of training images instead of all images from a set (3/1) of volunteers. The same trend is shown for the set n7 using models R and TL.

Using only end-systolic and end-diastolic images led to similar convergence speed and peak performance regarding DICE scores compared to the full data set (R_LV,MY_: 0.90, 0.77; TL_LV,MY_: 0.90, 0.78; TL^2^_LV,MY_: 0.92, 0.81 versus R_LV,MY_: 0.89, 0.77; TL _LV,MY_: 0.91, 0.79; TL^2^ _LV,MY_: 0.92, 0.81). In addition, the selection of end-systolic and end-diastolic images led to increased DICE-scores as starting points, fast convergence, and higher peak performance for all models, when compared to the same number of randomly selected images.

## Discussion

In this study, we successfully used a specialized, publicly available model^4^ to produce labels for a public data set of clinical 1.5 and 3T cardiac cine MRI, enabling access to more annotated data. Based on these labels we created a basic AI model, other researchers can use for their individual segmentation tasks. In addition, we applied transfer learning to segmentation of 7T human cine data, demonstrating that models based on these labels and a moderate amount of new domain data enable state-of-the-art segmentation results.

One of the obstacles to get started in deep learning based segmentation is the large amount of annotated data required to train an initial model. In this study we circumvent this problem by using the public Kaggle data set, to which we provide labels. The quality of these labels was evaluated using the volume information (end-systolic and end-diastolic volumes) included in the original Kaggle data set. Therefore, careful data curation had to be applied to avoid data inconsistencies (slice spacing, changes in image dimensions and image resolution, as well as missing slices) within individual patients. In addition, we found that label quality was connected to image orientation and image resolution. Scores (mean distance between labels and Kaggle “ground truth”), data curation scripts, as well as labels are provided in the online repository, enabling future use in other studies. We want to point out that label quality and accuracy was assessed via comparison to volume information only, with rare exceptions of visual confirmation. Thresholds of 5%, 10%, and 15% (deviation to the “ground truth”) for the subsets used in this study were chosen arbitrarily. With 54540, 162480, and 239350 images respectively, we assumed these three sets to provide the reasonable compromise between label accuracy and label quantity needed to assess data requirements in this specific transfer learning application.

Based on the now annotated data we trained initial segmentation models with varying architectures (ResNet34, ResNet50, VGG16), varying loss functions (cross-entropy, generalized DICE, focal loss), varying training sets (p5, p10, p15). The final model we selected was a ResNet34, using cross-entropy as a loss function, and the p5 set for training with an image resolution of 256×256. While we selected this model based on performance (mean distance to ground truth EF), overall impacts of parameter variations (3.64-4.06% mean distance to ground truth EF) were rather small. Similar to the use in this study, researchers or clinicians can use this model as a starting point for their respective transfer learning applications.

Considering the performance of this model on 7T human cine data (DICE_LV_: 0.84, DICE_MY_: 0.67), generalization capability appears limited. This is also true for the UKBB model (7T human cine, DICE_LV_: 0.67, DICE_MY_: 0.52). As the authors^4^ point out, the UKBB model was “trained on a single data set, the UK Biobank dataset, which is a relatively homogenous dataset” and might therefore “not generalize well to other vendor or sequence datasets”. With respect to the performance on 7T data this just means that, compared to the UKBB dataset, the Kaggle data set contains image patterns and characteristics more similar to the 7T data we acquired. In addition, it emphasizes why improvements in generalization^37-39^ are needed and why we applied an additional step of transfer learning to 7T data.

Due to differences in training data our initial models based on UKBB labels outperformed the UKBB model on the Kaggle data. While the UKBB model was trained on the homogeneous UKBB data, our models were trained on the heterogeneous Kaggle data itself. In addition, we applied data augmentation with respect to rotations and contrast and used only Kaggle data with the most accurate (top 15%) labels.

While multiple studies^4,5,26,40^ have demonstrated great image segmentation results for one specific dataset, these models have not been tested on other datasets or initially lack generalization capability. In this study, we show that transfer learning leads to improved model performance. DICE scores achieved on 7T human cine data prior to and after transfer learning were DICE_LV_: 0.84, DICE_MY_: 0.67 and DICE_LV_: 0.92, DICE_MY_: 0.81, respectively. This was comparable to human inter-observer variability (DICE_LV_: 0.94 and DICE_MY_: 0.81) and is within the range of state-of-the-art results, despite the relatively small set of training data^19^. In addition, inter-observer-variability in EDV (3.5%) and ESV (10.5%) between our model and the expert radiologist are in good agreement with literature reports (EDV: 2.5-5.3%, ESV: 6.8-13.9%)^41^ based on SSFP CMR imaging.

Typically, segmentation of the left ventricle is done to evaluate ejection fraction, a clinically used parameter. In this study we show that the model based volume prediction on the test set is very accurate for apical, mid-cavity and basal slices, with the exception of the most basal slice, where myocardial tissue moves in and out of plane throughout the cardiac cycle. Since we do not have a “ground-truth” segmentation for the Kaggle data and no information on labelling protocols, we do not know if there is any consistency in the definition of basal slices or the inclusion or exclusion of papillary muscle.

While transfer learning allows models to adapt to similar tasks and new datasets, containing new characteristics and patterns, this step also requires new labels. This aspect is often a limitation, since labelled medical data is difficult to acquire, particularly in areas that require domain-specific knowledge. In addition, the manual labelling process for high quality segmentations itself is often tedious and labor intensive. In this study we show that transfer learning applications (ImageNet weights to Kaggle data to 7T data) for cardiac cine segmentation of human 7T data can provide state-of-the-art results when training with labelled data from 7-14 volunteers (2626 – 5076 images), reaching DICE_LV_: 0.92 and DICE_MY_: 0.81 as well as accurate EF values. Having labels for three volunteers (1001 images) leads to decent results (DICE_LV_: 0.91 and DICE_MY_: 0.80). We consider labels for only one volunteer to be insufficient (DICE_LV_: 0.88 and DICE_MY_: 0.77).

For small training datasets (n≤1001) we show that a random selection of images from multiple volunteers leads to better performance compared to the selection of all images from a smaller number of volunteers (n=3 or n=1, figure 6). Generalization capabilities of a model increase with the amount of variation provided in the training data and thus using data from a multitude of patients or volunteers, where morphology and therefore image content and contrast differ, may be more beneficial than providing the same number of more coherent images from a small number of volunteers. Furthermore we demonstrate that the number of required images can drastically be reduced (from 5076 to 448 images), using labelled data from specific heart phases, end-diastolic and end-systolic, instead of all images. This may be possible, because knowing the two extreme states of contraction the model can deal more easily with intermediate states. Considering that n=448 images (roughly two cardiac EF examinations) enable close to state-of-the-art results for cardiac cine segmentation, data requirements for transfer learning applications in closely related tasks are low. In addition, labels for end-diastolic and end-systolic images are created in routine clinical cardiac examinations and thus easily accessible.

In summary, how much and which kind of data should be included in the transfer learning process should be carefully considered prior to labelling new data. In particular, the notion to provide data patient by patient may result in higher data requirements than necessary. There are various other routine cardiac MR examinations such as T_2_, T_1_, LGE, and even T_2_^*^ that require segmentation^38,39,42^. Transfer learning applications to image segmentation of such varying contrasts may benefit from the amount of annotated data and the framework provided in this study.

With respect to future use of this annotated data we recommend researchers take the following steps:

1. use the pre-trained model we provide (r34_CE_p5_s2)
2. re-train with training data from the new domain and tune hyper parameters using validation data from the new domain
3. evaluate model performance on a test set from the new domain

In this study, we used only the 5-15% of the most accurate kaggle labels to create our base models. Thus, researchers attempting to train their own base network using the labelled Kaggle data should always assess label quality.

The experimental 7T data used in this study is not comparable to clinical cardiac MRI in patients. Future performances on clinical data should be evaluated against the Kaggle dataset. There are some limitations connected to the use of the Kaggle dataset. While there are variations in measurement parameters, such as resolution, FOV, matrix size, TE, TR, bandwidth, and slice thickness, most examinations (∼90%) were done at 1.5T. In addition, all data was acquired using Siemens whole body MRI systems. Models trained using this dataset might thus not generalize well to other vendor datasets, requiring transfer learning as demonstrated in this study.

Since no disease-related information is provided in the Kaggle dataset, we have no knowledge which and how many pathological patterns are currently represented in the dataset. In this study we demonstrate that transfer learning to 7T data of healthy human volunteers enables DICE scores of DICE_LV_: 0.92 and DICE_MY_: 0.81. A clinical application would require a performance assessment or transfer learning for specific cardiac pathologies, both beyond the scope of this cardiology-related methodological work.

Furthermore, the accuracy of the labels we created was assessed based on comparison to provided volume information only and visual confirmation of the contours may be biased, because we do not know if the provided volume information is based on consistent definitions of basal slices or the inclusion or exclusion of papillary muscle. This should be considered when creating models based on this dataset. In general, there is a need for a standard benchmark dataset, where labels are based on standardized protocols and images are representations of diverse clinical phenotypes (diseases, vendors, field strengths, sequences, protocols).

### Conclusions

In this study, we provide access to annotated cardiac cine MRI data, and AI models, which can be used as a starting point for transfer learning applications. Using such a base model, we demonstrate that transfer learning from clinical 1.5 and 3T cine data to 7T cine data is feasible with moderate data requirements, enabling future applications to other cardiac MRI examinations such as T_2_, T_1_, LGE, and even T_2_^*^. Furthermore, we show that not all data has the same value with respect to transfer learning approaches and that careful selection of the training data may drastically reduce data requirements.

## Data Availability

Data used and generated in this study is available at Zenodo. Source code, models and detailed instructions for reproducibility are available at GitHub and archived at Zenodo. Due to data privacy regulations the 7T MR images are not publically available.

https://doi.org/10.5281/zenodo.3876351

https://github.com/chfc-cmi/cmr-seg-tl

https://doi.org/10.5281/zenodo.3894646

## Declarations

### Ethics approval and consent to participate

Ethics approval of the local ethics committee at the University Hospital Würzburg has been granted under reference number 7/17-SC.

### Consent for publication

All human volunteers gave their consent for publication using our institutional consent form.

### Availability of data and materials

The datasets supporting the conclusions of this article are available in the zenodo repository, https://doi.org/10.5281/zenodo.3876350 and source code is available in GitHub and zenodo https://github.com/chfc-cmi/cmr-seg-tl, https://doi.org/10.5281/zenodo.3894647. The raw 7T images are not publicly available due to data privacy regulations.

### Authors’ contributions

DL, MJA, LMS & TW designed the study. MJA, DL & TW developed the computational methods. DL, WS and TR collected the data. MJA and DL analyzed and interpreted the data. DL wrote the initial draft of the manuscript. All authors read and approved the final manuscript.

## Acknowledgements

We thank Andreas Hotho for insightful discussions.

## List of abbreviations

LV: left ventricle
RV: right ventricle
EF: ejection fraction
ESV: end-systolic volume
EDV: end-diastolic volume
lr: learning rate
UKBB: UK Biobank

## References

1. Moon JC, Lorenz CH, Francis JM, Smith GC, Pennell DJ. Breath-hold FLASH and FISP cardiovascular MR imaging: left ventricular volume differences and reproducibility. Radiology. Jun 2002;223(3):789–797.

2. Curtis JP, Sokol SI, Wang Y, et al. The association of left ventricular ejection fraction, mortality, and cause of death in stable outpatients with heart failure. J Am Coll Cardiol. Aug 20 2003;42(4):736–742.

3. Karamitsos TD, Francis JM, Myerson S, Selvanayagam JB, Neubauer S. The role of cardiovascular magnetic resonance imaging in heart failure. J Am Coll Cardiol. Oct 6 2009;54(15):1407–1424.

4. Bai W, Sinclair M, Tarroni G, et al. Automated cardiovascular magnetic resonance image analysis with fully convolutional networks. Journal of cardiovascular magnetic resonance : official journal of the Society for Cardiovascular Magnetic Resonance. Sep 14 2018;20(1):65.

5. Baumgartner CF, Koch LM, Pollefeys M, Konukoglu E. An Exploration of 2D and 3D Deep Learning Techniques for Cardiac MR Image Segmentation. arXiv e-prints. 2017. https://ui.adsabs.harvard.edu/abs/2017arXiv170904496B. Accessed September 01, 2017.

6. Jang Y, Hong Y, ha S, Kim S, Chang H-J. Automatic Segmentation of LV and RV in Cardiac MRI. 2018:161–169.

7. Tran PV. A Fully Convolutional Neural Network for Cardiac Segmentation in Short-Axis MRI. arXiv e-prints. 2016. https://ui.adsabs.harvard.edu/abs/2016arXiv160400494T. Accessed April 01, 2016.

8. Liu J, Pan Y, Li M, et al. Applications of deep learning to MRI images: A survey. Big Data Mining and Analytics. 2018;1(1):1–18.

9. Chen F, Taviani V, Malkiel I, et al. Variable-Density Single-Shot Fast Spin-Echo MRI with Deep Learning Reconstruction by Using Variational Networks. Radiology. Nov 2018;289(2):366–373.

10. Schlemper J, Caballero J, Hajnal JV, Price AN, Rueckert D. A Deep Cascade of Convolutional Neural Networks for Dynamic MR Image Reconstruction. IEEE transactions on medical imaging. Feb 2018;37(2):491–503.

11. Zhu B, Liu JZ, Cauley SF, Rosen BR, Rosen MS. Image reconstruction by domaintransform manifold learning. Nature. 2018/03/01 2018;555(7697):487–492.

12. Benou A, Veksler R, Friedman A, Riklin Raviv T. Ensemble of expert deep neural networks for spatio-temporal denoising of contrast-enhanced MRI sequences. Med Image Anal. Dec 2017;42:145–159.

13. Bermudez C, Plassard AJ, Davis TL, Newton AT, Resnick SM, Landman BA. Learning Implicit Brain MRI Manifolds with Deep Learning. Proceedings of SPIE--the International Society for Optical Engineering. Mar 2018;10574.

14. de Vos BD, Berendsen FF, Viergever MA, Sokooti H, Staring M, Isgum I. A Deep Learning Framework for Unsupervised Affine and Deformable Image Registration. arXiv e-prints. 2018. https://ui.adsabs.harvard.edu/abs/2018arXiv180906130D. Accessed September 01, 2018.

15. Wu G, Kim M, Wang Q, Munsell BC, Shen D. Scalable High-Performance Image Registration Framework by Unsupervised Deep Feature Representations Learning. IEEE transactions on bio-medical engineering. Jul 2016;63(7):1505–1516.

16. Akkus Z, Galimzianova A, Hoogi A, Rubin DL, Erickson BJ. Deep Learning for Brain MRI Segmentation: State of the Art and Future Directions. Journal of Digital Imaging. 2017/08/01 2017;30(4):449–459.

17. Hesamian MH, Jia W, He X, Kennedy P. Deep Learning Techniques for Medical Image Segmentation: Achievements and Challenges. Journal of Digital Imaging. 2019/08/01 2019;32(4):582–596.

18. Ruijsink B, Puyol-Antón E, Oksuz I, et al. Fully Automated, Quality-Controlled Cardiac Analysis From CMR: Validation and Large-Scale Application to Characterize Cardiac Function. JACC: Cardiovascular Imaging. 2019/07/17/ 2019.

19. Chen C, Qin C, Qiu H, et al. Deep learning for cardiac image segmentation: A review. arXiv e-prints. 2019. https://ui.adsabs.harvard.edu/abs/2019arXiv191103723C. Accessed November 01, 2019.

20. Liu F, Shen C. Learning Deep Convolutional Features for MRI Based Alzheimer’s Disease Classification. arXiv e-prints. 2014. https://ui.adsabs.harvard.edu/abs/2014arXiv1404.3366L. Accessed April 01, 2014.

21. Pinaya WHL, Gadelha A, Doyle OM, et al. Using deep belief network modelling to characterize differences in brain morphometry in schizophrenia. Scientific Reports. 2016/12/12 2016;6(1):38897.

22. Bello GA, Dawes TJW, Duan J, et al. Deep-learning cardiac motion analysis for human survival prediction. Nature Machine Intelligence. 2019/02/01 2019;1(2):95–104.

23. Dawes TJW, de Marvao A, Shi W, et al. Machine Learning of Three-dimensional Right Ventricular Motion Enables Outcome Prediction in Pulmonary Hypertension: A Cardiac MR Imaging Study. Radiology. May 2017;283(2):381–390.

24. Lundervold AS, Lundervold A. An overview of deep learning in medical imaging focusing on MRI. Zeitschrift für Medizinische Physik. 2019/05/01/ 2019;29(2):102–127.

25. Sudlow C, Gallacher J, Allen N, et al. UK Biobank: An Open Access Resource for Identifying the Causes of a Wide Range of Complex Diseases of Middle and Old Age. PLOS Medicine. 2015;12(3):e1001779.

26. Data Science Bowl Cardiac Challenge Data 2016. https://www.kaggle.com/c/second-annual-data-science-bowl/data. Accessed 29th of July 2019.

27. Paszke A, Gross S, Massa F, et al. PyTorch: An Imperative Style, High-Performance Deep Learning Library. arXiv e-prints. 2019:arXiv:1912.01703. https://ui.adsabs.harvard.edu/abs/2019arXiv191201703P. Accessed December 01, 2019.

28. Howard J, Gugger S. Fastai: A Layered API for Deep Learning. Information. 2020;11(2):108.

29. Ronneberger O, Fischer P, Brox T. U-Net: Convolutional Networks for Biomedical Image Segmentation. arXiv e-prints. 2015:arXiv:1505.04597. https://ui.adsabs.harvard.edu/abs/2015arXiv150504597R. Accessed May 01, 2015.

30. He K, Zhang X, Ren S, Sun J. Deep Residual Learning for Image Recognition. arXiv e-prints. 2015:arXiv:1512.03385. https://ui.adsabs.harvard.edu/abs/2015arXiv151203385H. Accessed December 01, 2015.

31. Abbasi-Sureshjani S, Amirrajab S, Lorenz C, Weese J, Pluim J, Breeuwer M. 4D Semantic Cardiac Magnetic Resonance Image Synthesis on XCAT Anatomical Model. arXiv e-prints. 2020. https://ui.adsabs.harvard.edu/abs/2020arXiv200207089A. Accessed February 01, 2020.

32. Simonyan K, Zisserman A. Very Deep Convolutional Networks for Large-Scale Image Recognition. arXiv e-prints. 2014:arXiv:1409.1556. https://ui.adsabs.harvard.edu/abs/2014arXiv1409.1556S. Accessed September 01, 2014.

33. Smith LN. A disciplined approach to neural network hyper-parameters: Part 1 --learning rate, batch size, momentum, and weight decay. arXiv e-prints. 2018:arXiv:1803.09820. https://ui.adsabs.harvard.edu/abs/2018arXiv180309820S. Accessed March 01, 2018.

34. Sudre CH, Li W, Vercauteren T, Ourselin S, Jorge Cardoso M. Generalised Dice Overlap as a Deep Learning Loss Function for Highly Unbalanced Segmentations. 2017; Cham.

35. Lohr D, Terekhov M, Kosmala A, Stefanescu MR, Hock M, Schreiber LM. Cardiac MRI with the Siemens Terra 7T System: Initial Experience and Optimization of Default Protocols. Paper presented at: Proc. of the 26th Annual Meeting of ISMRM; April, 2018; Paris, France.

36. Zijdenbos AP, Dawant BM, Margolin RA, Palmer AC. Morphometric analysis of white matter lesions in MR images: method and validation. IEEE transactions on medical imaging. 1994;13(4):716–724.

37. Feng X, Yang J, Laine AF, Angelini ED. Discriminative Localization in CNNs for Weakly-Supervised Segmentation of Pulmonary Nodules. arXiv e-prints. 2017:arXiv:1707.01086. https://ui.adsabs.harvard.edu/abs/2017arXiv170701086F. Accessed July 01, 2017.

38. Chen J, Li H, Zhang J, Menze B. Adversarial Convolutional Networks with Weak Domain-Transfer for Multi-Sequence Cardiac MR Images Segmentation. arXiv e-prints. 2019. https://ui.adsabs.harvard.edu/abs/2019arXiv190809298C. Accessed August 01, 2019.

39. Wang J, Huang H, Chen C, Ma W, Huang Y, Ding X. Multi-sequence Cardiac MR Segmentation with Adversarial Domain Adaptation Network. arXiv e-prints. 2019. https://ui.adsabs.harvard.edu/abs/2019arXiv191012514W. Accessed October 01, 2019.

40. Tran PV. A Fully Convolutional Neural Network for Cardiac Segmentation in Short-Axis MRI. arXiv e-prints. 2016:arXiv:1604.00494. https://ui.adsabs.harvard.edu/abs/2016arXiv160400494T. Accessed April 01, 2016.

41. Luijnenburg SE, Robbers-Visser D, Moelker A, Vliegen HW, Mulder BJM, Helbing WA. Intra-observer and interobserver variability of biventricular function, volumes and mass in patients with congenital heart disease measured by CMR imaging. Int J Cardiovasc Imaging. 2010;26(1):57–64.

42. Vesal S, Ravikumar N, Maier A. Automated Multi-sequence Cardiac MRI Segmentation Using Supervised Domain Adaptation. arXiv e-prints. 2019. https://ui.adsabs.harvard.edu/abs/2019arXiv190807726V. Accessed August 01, 2019.

